# Cytokine profile in plasma of severe COVID-19 does not differ from ARDS and sepsis

**DOI:** 10.1101/2020.05.15.20103549

**Authors:** Jennifer G. Wilson, Laura J. Simpson, Anne-Maud Ferreira, Arjun Rustagi, Jonasel Roque, Adijat Asuni, Thanmayi Ranganath, Philip M. Grant, Aruna Subramanian, Yael Rosenberg-Hasson, Holden T. Maecker, Susan P. Holmes, Joseph E. Levitt, Catherine A. Blish, Angela J. Rogers

## Abstract

**Rationale:** Elevated levels of inflammatory cytokines have been associated with poor outcomes among COVID–19 patients. It is unknown, however, how these levels compare to those observed in critically ill patients with ARDS or sepsis due to other causes.

**Objectives:** To directly compare plasma levels of inflammatory cytokines, with a focus on 6 cytokines associated with cytokine storm (IL-1b, IL-1RA, IL-6, IL-8, IL-18, and TNFα), between hospitalized COVID-19 patients and banked plasma samples from ARDS and sepsis patients from prior to the COVID-19 pandemic.

**Findings:** 15 hospitalized COVID-19 patients, 9 of whom were critically ill, were compared to 28 critically ill patients with ARDS or sepsis. There were no statistically significant differences in baseline levels of IL-1b, IL-1RA, IL-6, IL-8, IL-18, and TNFα between patients with severe COVID-19 and critically ill controls with ARDS or sepsis.

**Conclusions:** Levels of inflammatory cytokines IL-1b, IL-1RA, IL-6, IL-8, IL-18, and TNFα were not higher in critically ill COVID-19 patients than in critically ill patients admitted with ARDS or sepsis due to other causes in this small cohort. Broad use of immunosuppressive therapies in ARDS has failed in numerous Phase 3 studies; use of these therapies in unselected patients with COVID-19 is likely unwarranted.

## INTRODUCTION

Numerous reports have described an association between elevated inflammatory markers and poor outcomes in COVID-19 patients.^1–3^ These data have sparked interest in “cytokine storm” as a major driver of illness severity in COVID-19, and multiple clinical trials are underway to test the efficacy of immunosuppressive therapies, including IL-6 antagonists. However, it is unclear if inflammatory cytokine levels are truly higher in patients with severe COVID-19 than in critically ill patients with ARDS or sepsis due to other causes.

While ARDS and sepsis are recognized to be highly inflamed states, large-scale randomized controlled trials of immune modulators (e.g. anti-IL1b, activated protein C, steroids) have failed.^4,5^ Recent reporting of IL-6 levels in severe COVID-19 patients (∼10–40 pg/mL measured in clinical labs)^1–3^ are lower than levels reported in prior ARDS cohorts (∼100–2000 pg/mL using ELISA).^6–8^ Given potential variability in cytokine quantification across platforms, we measured plasma levels of 76 cytokines, including six inflammatory cytokines associated with cytokine storm (IL-1b, IL-1RA, IL-6, IL-8, IL-18, and TNFα), in a prospective cohort of patients hospitalized with COVID-19 and directly compared them to levels in banked biospecimens from ARDS and sepsis patients enrolled in the Stanford ICU Biobank prior to the COVID-19 pandemic.

## RESULTS AND DISCUSSION

Clinical characteristics of the study participants in each group are shown in **Table 1**. Of the 9 severe COVID-19 patients, 6 required mechanical ventilation. While only 1 ventilated COVID-19 patient died, 5 had moderate-to-severe ARDS (P:F<150 at enrollment) and all required prolonged mechanical ventilation (>7 days).

**Table 1.** Patient Data by Study GroupIQR = interquartile range. ARDS = acute respiratory distress syndrome. SOFA =Sequential Organ Failure Assessment (SOFA) score, which ranges from 0 to 24,withhigher scores indicating more severe organ failure.

| **Table 1. Patient Data by Study Group** |  |  |  |
| --- | --- | --- | --- |
|  | **Non-ICU COVID-19**<br>**(N = 6)** | **ICU COVID-19**<br>**(N = 9)** | **Sepsis/ARDS**<br>**(N = 28)** |
| Age, median (IQR), yrs | 53 (43, 62) | 58 (40, 64) | 58 (34, 69) |
| Male, *n* (%) | 5 (83%) | 6 (67%) | 19 (68%) |
| Hispanic ethnicity, *n* (%) | 0% | 4 (44%) | 4 (14%) |
| White race, *n* (%) | 4 (67%) | 6 (66%) | 20 (71%) |
| Active malignancy, *n* (%) | 0% | 1 (11%) | 3 (14.%) |
| Immunosuppression, *n* (%) | 0% | 1 (11%) | 5 (18%) |
| SOFA at enrollment, median (IQR) | 2 (1, 2) | 8 (2, 10) | 8 (6, 10) |
| Vasopressors, *n* (%) | n/a | 6 (67%) | 23 (82%) |
| ARDS criteria, *n* (%) | 0% | 6 (67%) | 12 (43%) |
| Mechanically ventilated, *n* (%) | n/a | 6 (67%) | 13 (46%) |
| Died prior to hospital discharge, *n* (%), | 0% | 1 (11%) | 6 (21%) |

Levels of IL-1b, IL-1RA, IL-6, IL-8, IL-18 and TNFα did not significantly differ between the moderate COVID-19, severe COVID-19, and ARDS/sepsis group (**Figure 1**, after adjustment for multiple comparisons, all p values > 0.05). There was a trend towards higher levels of IL-1RA and IL-6 in the patients with severe COVID-19 as compared to those with moderate COVID-19, consistent with prior reports.^1–3^ There was also a trend towards higher IL-18 in the severe COVID-19 group compared to the other critical illness group (p-unadjusted = 0.01, p-adjusted = 0.10).

**Figure 1.**
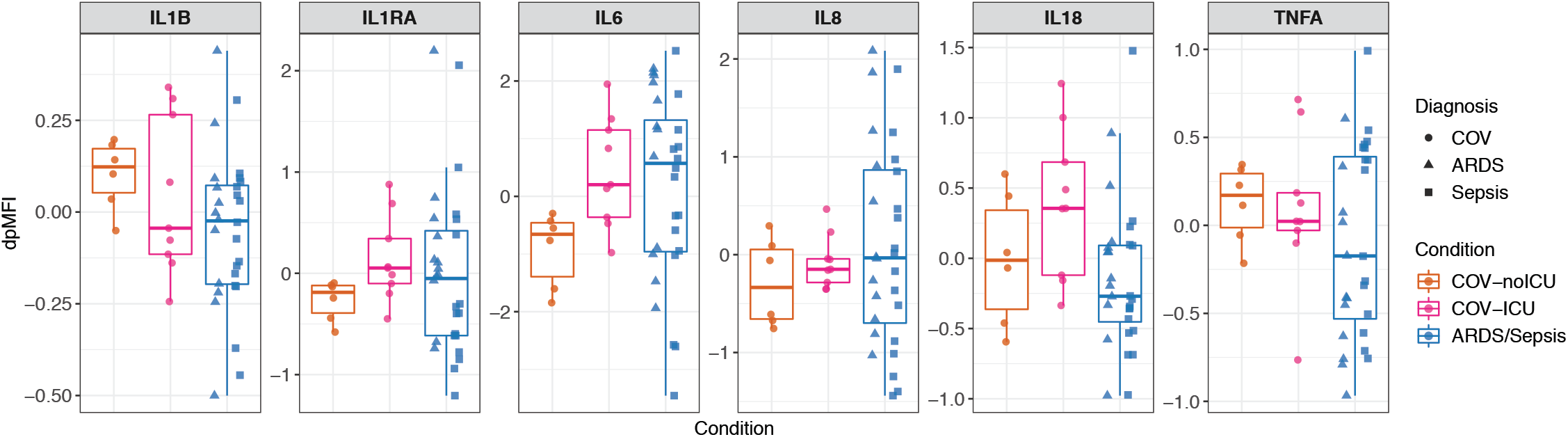
Expression of inflammatory cytokines does not vary between COVID ICU patients and samples from non-COVID sepsis and ARDS. Cytokine expression levels of two replicates per sample were measured in plasma using Luminex, and normalized for nonspecific binding, resulting in the mean of plate-detrended median fluorescence intensity (dpMFI) values for each cytokine per sample. Moderate COVID-19 (COV-noICU,orange), n = 6. Severe COVID-19 (COV-ICU, pink), n = 9. ARDS/Sepsis (blue), n = 28. Circles represent subjects with COVID-19, triangles represent subjects with ARDS, and squares represent subjects with sepsis. Wilcoxon rank sum tests were performed for each cytokine, comparing COV-ICU to COV-noICU, and COV-ICU to ARDS/Sepsis. P-values were adjusted for multiple comparisons using the Benjamini-Hochberg correction method. No cytokines had adjusted p-values < 0.05. P-values for comparison between COV-noICU and COV-ICU: IL1B, p = 0.53, p-adjusted = 0.70; IL1RA, p = 0.035, p-adjusted = 0.14; IL6, p = 0.018, p-adjusted = 0.11; IL8, p = 0.46, p-adjusted = 0.68; IL18, p = 0.27, p-adjusted = 0.68; TNFα, p = 0.86, p-adjusted = 0.90. P-values for comparison between COV-ICU and ARDS/Sepsis: IL1B, p = 0.41, p-adjusted = 0.68; IL1RA, p = 0.35, p-adjusted = 0.68; IL6, p = 0.90, p-adjusted = 0.90; IL8, p = 0.85, p-adjusted = 0.90; IL18, p = 0.01, p-adjusted = 0.10; TNFA, p = 0.37, p-adjusted = 0.68.

In the more extended exploratory analysis of 70 additional cytokines, the three patient groups did not differ strongly in principal component analysis (**Supplemental Figure S1**, available in the online supplement). There were small but statistically significant differences in the levels of IL-16, IL-21, IL-28A (IFNL2), and TSLP between the three groups (**Supplemental Figure S2**), but no cytokine levels were dramatically increased in COVID-19 compared to other causes of critical illness (**Supplemental Figures S2** and **S3**). Together, these data suggest that a “cytokine storm” in COVID-19 that is distinct from other critical illness (e.g. sepsis and ARDS) is unlikely.

Our primary goal was to directly compare levels of six inflammatory cytokines commonly associated with cytokine storm (IL-1b, IL-1RA, IL-6, IL-8, IL-18, and TNFα) between severe COVID-19 patients and patients with ARDS or sepsis due to other causes. Our findings are consistent with previous data demonstrating higher levels of inflammatory cytokines among COVID-19 patients with more severe disease. Importantly, however, our data suggest that inflammatory biomarkers in severe COVID-19 patients are *not* markedly elevated when directly compared to critically ill patients with ARDS or sepsis.

IL-6 levels were measured by the clinical lab (as part of clinical care) in 6 of the COVID-19 patients, including 4 with severe disease, though not at matched time points with the research blood collection. Levels ranged from <6–31 pg/mL. Given the small number of patients who had clinical measurements available, and the variation in collection times, we were unable to derive accurate concentrations based on MFI for the remaining patients. Nonetheless, these data points further support our findings that IL-6 levels in particular—while elevated above levels found in healthy subjects—are not markedly elevated in all severe COVID-19 patients compared to other critically ill patients.

This brief report calls into question the idea that “cytokine storm” is the major driver of morbidity and mortality in all severe COVID-19 patients. As Ritchie and Singanayagam have stated, it is equally possible that the higher levels of proinflammatory cytokines seen in severe COVID-19 reflect an increased burden of virus rather than “an inappropriate host response that requires correction.”^9^

The most important limitation of this study is our small sample size. Even in our predetermined analysis of six cytokines strongly associated with cytokine storm, we lack power to detect minor differences between groups. The exploratory analyses of an additional 70 cytokines is similarly limited, but is provided as a reference for the field. In addition, we do not have measurements of cytokines over time, but only near the point of enrollment. Finally, as discussed above, we report cytokine levels by MFI per recommended Luminex analysis methods,^10^ precluding direct comparison of our values to previously published data that report cytokine levels in pg/mL.

Despite the above limitations, it is clear that in this cohort, inflammatory cytokines are not dramatically higher in severe COVID-19 patients than in other critically ill patients. Broad testing of immunosuppressive therapies in unselected COVID-19 patients may therefore face the same fate as trials of immunosuppression in unselected ARDS and sepsis patients: inconclusive results despite decades of effort, and the uncomfortable understanding that while some patients may have been helped, others may have been harmed. Indeed, given the duration of mechanical ventilation and attendant high rates of nosocomial infection in severe COVID-19 patients, these therapies have potential for harm. Further research is needed to characterize the spectrum of the immune response in COVID-19, and to identify which patients are most likely to benefit from (and least likely to be harmed by) immunomodulatory therapies.

## METHODS

Sequential inpatients with COVID-19 admitted to Stanford Hospital were approached for participation in the Stanford ICU biobank between mid-March and early April, 2020. Patients who were enrolled in randomized clinical trials that required blood draws were excluded. COVID-19 patients were classified as “severe” if admitted to the Intensive Care Unit (ICU), and “moderate” if they did not require ICU admission.

Twenty-eight critically ill subjects with ARDS and sepsis who had been enrolled in the Stanford ICU biobank between 2015–2018 were selected for comparison. Briefly, the Stanford ICU Biobank recruits patients at risk for development of respiratory failure and ARDS admitted to Stanford Hospital as previously described.^11^ Subjects are eligible for enrollment when decision to admit to ICU is made, either from the Emergency Department or the hospital wards, with goal enrollment in <24h of ICU transfer. All 28 patients were phenotyped for ARDS and sepsis by 2-physician consensus (AJR and JEL), based on the Berlin Criteria and Sepsis-2 criteria and using all available hospital clinical data including history, physical exam, laboratory and microbiologic data, invasive monitoring data, autopsy results, and physician summaries. The Stanford ICU is a major referral center for cancer and thus typically has high rates of immunosuppression. To assess cytokine response to infection in patients with a normal immune system (similar to the COVID population), the ARDS and sepsis patients were therefore enriched for normal baseline immune system (e.g. no metastatic cancer, bone marrow transplant, or high dose steroids) in comparison to the Biobank as a whole.

Luminex assays were performed at the Human Immune Monitoring Center at Stanford University. All samples were run in duplicate in a single plate per panel. Kits were purchased from EMD Millipore Corporation, Burlington, MA., and used according to the manufacturer’s recommendations. Custom Assay Chex control beads were purchased from Radix Biosolutions, Georgetown, Texas,and were added to all wells and their median fluorescence intensity (MFI) was quantified and used to assess and normalize for non-specific binding.

Full details on cytokine measurement technique are available as an appendix in the online supplement.

### Statistics

All statistical analyses were performed using the open source statistical software R (https://www.r-project.org). Because we observed significant differences in CHEX4 MFI between the three groups tested, we corrected for CHEX4 nonspecific binding, keeping the clinical conditions (severe COVID-19 (COV-ICU), moderate COVID-19 (COV-noICU), ARDS and Sepsis), age, and sex as covariates. Each sample was normalized according to methods used by the Human Immune Monitoring Center at Stanford University.^12^ Briefly, the median fluorescence intensity (MFI) of each cytokine was corrected first for plate/batch/lot artifacts by linear mixed modeling, then the average of technical replicates was log transformed. Then the log transformed average MFIs were corrected for nonspecific binding by local polynomial regression and repeated cross-validation, resulting in plate-detrended MFI (dpMFI) values.

Once normalized for non-specific binding, levels of IL-1b, IL-1RA, IL-6, IL-8, IL-18, and TNFα in severe COVID-19 patients (COV-ICU) were compared to levels in moderate COVID-19 patients (COV-noICU) and to levels in patients with other critical illness (ARDS and sepsis) using the Wilcoxon rank sum test corrected for multiple testing by the Benjamini-Hochberg method. Kruskal-wallis tests were performed on the remaining 70 cytokines, and p-values were corrected for multiple testing by the Benjamini-Hochberg method.

### Study approval

This study was conducted according to Declaration of Helsinki principles, and was approved by the Stanford University Hospital IRB (protocol 28205). All patients or their surrogates gave written informed consent to participate in the Stanford ICU Biobank.

## Data Availability

The data will be made available at ImmPort (immport.org) upon publication of this manuscript in a peer-reviewed journal.

## Author Contributions

JGW, LJS, AMF, CAB, and AJR participated in project conception, design, and data interpretation. AR, JR, AA, TR, PMG, AS, JEL, AJR contributed to patient enrollment, data acquisition and sample processing. Data analysis was performed by LJS, AMF, YRH, HTM, SPH, and CAB. The manuscript was drafted by JGW, LJS, AMF, CAB, and AJR, with critical revisions by AR, PMG, AS, YRH, and JEL. All authors approved the final version of this article.JGW, LJS, and AMF made equal and critical contributions; JGW is listed first for her role in the conception of the study; LJS and AMF played a critical role in data analysis and interpretation.

## Acknowledgements

The authors thank all of the patients, families and care providers who supported this work.

## Conflict of interest statement

The authors declare that no conflict of interest exists.

## Notes

### Competing Interest Statement

The authors have declared no competing interest.

### Funding Statement

The Stanford ICU Biobank is funded by Dr. Rogers’ NHLBI K23
HL125663. C.A.B. is supported by the Burroughs Wellcome Fund Investigators in the Pathogenesis of Infectious Diseases #1016687, NIH/NIAID U19AI057229-16 (PI MM Davis), and is the Tashia and John Morgridge Faculty Scholar in Pediatric Translational Medicine from the Stanford Maternal Child Health Research Institute and an Investigator of the Chan Zuckerberg Biohub.

